# Peripheral blood DNA methylation signatures predict response to vedolizumab and ustekinumab in adult patients with Crohn’s disease: The EPIC-CD study

**DOI:** 10.1101/2024.07.25.24310949

**Authors:** Vincent W. Joustra, Andrew Y.F. Li Yim, Peter Henneman, Ishtu Hageman, Tristan de Waard, Evgeni Levin, Alexandra J. Noble, Thomas P. Chapman, Femke Mol, Sarah van Zon, Donghyeok Lee, Colleen G.C. McGregor, Alex T. Adams, Jack J. Satsangi, Wouter J. de Jonge, Geert R. D’Haens, EPIC-CD Consortium

## Abstract

Biological therapeutics are now widely used in Crohn’s disease (CD), with evidence of efficacy from randomized trials and real-world experience. Primary non-response is a common, poorly understood problem. We assessed blood methylation as a predictor of response to vedolizumab (VDZ, anti-a4b7 integrin) or ustekinumab (USTE, anti-IL-12/23p40). We report a two-center, prospective cohort study in which we profiled the peripheral blood DNA methylome of 184 adult male and female CD patients prior to and during treatment with VDZ or USTE in a discovery (n=126) and an external validation cohort (n=58). We defined epigenetic biomarkers that were stable over time and associated with combined clinical and endoscopic response to VDZ or USTE with an area under curve (AUC) of 0.87 and 0.89, respectively. We validated these models in an external cohort yielding an AUC of 0.75 for both VDZ and USTE. These data will now be prospectively tested in a multicenter randomized clinical trial.

**Graphical abstract:** 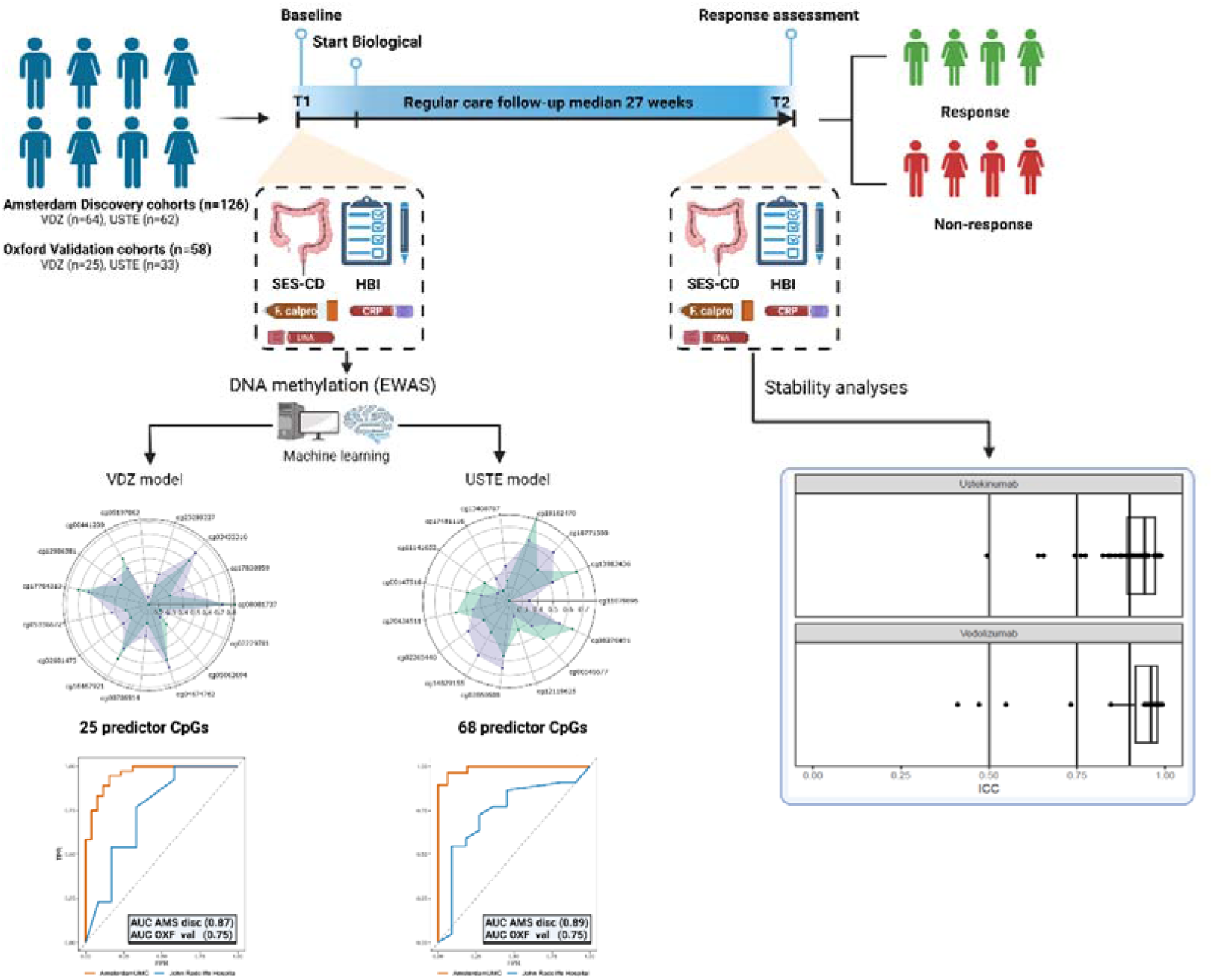

## Introduction

Crohn’s disease (CD) is an incurable, chronic, relapsing inflammatory bowel disease (IBD) caused by a complex interplay between the environment, gut microbiome, and a dysregulated immune system in genetically susceptible patients^1,2^. Accessibility to high-throughput “omics” technology has enhanced our understanding of the underlying molecular pathogenesis of CD leading to the development of several monoclonal antibodies or biologicals that target specific inflammatory pathways in an effort to suppress the inflammation and to induce and or maintain a state of clinical and endoscopic remission^3^. Currently, the repertoire of approved biologicals in CD includes anti-TNF antibodies (infliximab (IFX) and adalimumab (ADA)), the anti-α_4_β_7_ integrin antibody vedolizumab (VDZ) and the anti-IL12/23p40 antibody ustekinumab (USTE). More recently, specific IL23p19 antibodies and JAK inhibitors have also been approved for clinical use^4–6^. Despite the established efficacy of these biological treatments to induce corticosteroid-free *clinical remission* in up to 65% of CD patients, sustained *endoscopic remission* is observed in not more than a third of patients after 1 year of treatment^7–9^. This creates a clinical challenge since therapeutic guidelines suggest to use endoscopic remission as a target. An increasingly common clinical scenario is the choice between VDZ or USTE as second-line treatment for patients who have not responded to anti-TNF therapies.

To date, treatment selection has been based on a trial-and-error approach. Given the limited efficacy, many patients are therefore treated with insufficiently effective treatment, which is associated with an increased risk of complications (stenosis, fistula, abscesses, nutritional deficiencies) and surgery. The development of strategies that allow selection of treatment based on the likelihood of response is an important unmet need. Although various efforts using clinical^10^, transcriptomic^11–13^, proteomic^14^ or microbial^15,16^ technologies have been investigated and reported, no predictive biomarkers have made their way to clinical application^17–19^.

DNA methylation is one of the most studied epigenetic features characterized by the covalent binding of methyl groups to nucleotides, most often a cytosine in a cytosine-phosphate-guanine (CpG) sequence in humans^20^. DNA methylation is believed to play an essential role in the regulation of gene expression, thereby determining cellular phenotype and behavior without altering the DNA sequence itself^21,22^. Within the context of IBD, DNA methylation has gained particular interest due to its dynamic interaction with the environment and suggested epigenetic-microbial crosstalk^23–25^. A number of recent studies demonstrated differential DNA methylation profiles associated with the presence of CD and/or specific CD-phenotypes^26,27^ in peripheral blood leukocytes (PBL)^28–30^, intestinal mucosa^31^ and specific cell-types thereof^32,33^. Most studies proposed a potential role of the DNA methylome in diagnostics and prediction of treatment response.

In the current study we performed an epigenome-wide association study (EWAS) in which we identified and validated prognostic DNA methylation signatures in peripheral blood of adult CD patients associated with objective therapeutic response to VDZ and USTE. We subsequently interrogated whether these signatures were stable over time and assessed the influence of common confounding variables. We further performed additional validation analyses of the model against patients with previous non-response to one or both drugs. Finally, we investigated the association of the identified DNA methylation biomarkers with gene expression through transcriptomic analyses.

## Results

### Study population

We prospectively recruited a discovery cohort of 126 adult patients at the IBD Center of Amsterdam University Medical Centers (Amsterdam UMC), Amsterdam, Netherlands. All patients had active symptomatic and endoscopic CD and were scheduled to start VDZ (N = 64) or USTE (N = 62). Evidence for active disease was documented with a validated clinical score (Harvey Bradshaw Index (HBI), median 8 (interquartile range (IQR) 4-12)), biochemical tests (serum C-reactive protein (CRP), median 6.1 mg/L (IQR 2.2-14.8) and fecal calprotectin (FCP), median 903 µg/g (IQR 278-1816)) and also endoscopic signs of inflammation measured with the simple endoscopic score for CD (SES-CD), median 9 (IQR 6-15). In addition, an external validation cohort of 58 adult CD patients starting VDZ (N = 25) or USTE (N = 33) biological therapy were recruited at the John Radcliffe Hospital, Oxford, United Kingdom. Peripheral blood leukocyte (PBL) samples were obtained prior to treatment initiation and upon response assessment at a median of 27 (IQR 20-33) weeks into treatment, when patients were classified as responder (R) or non-responder (NR). A detailed overview of all the clinical characteristics across the different cohorts and treatments can be found in the methods section as well as in **Table 1**.

**Table 1:**
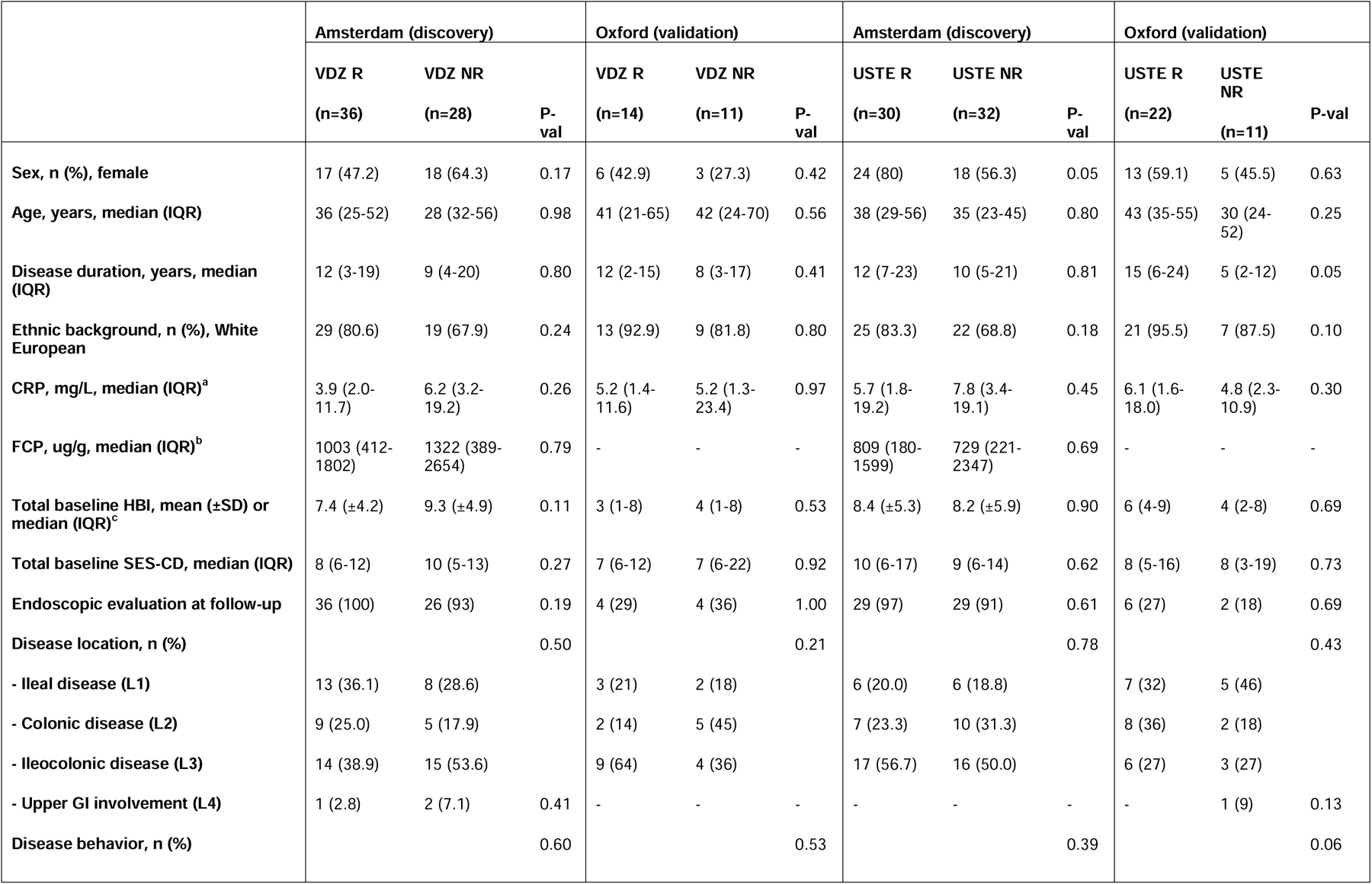
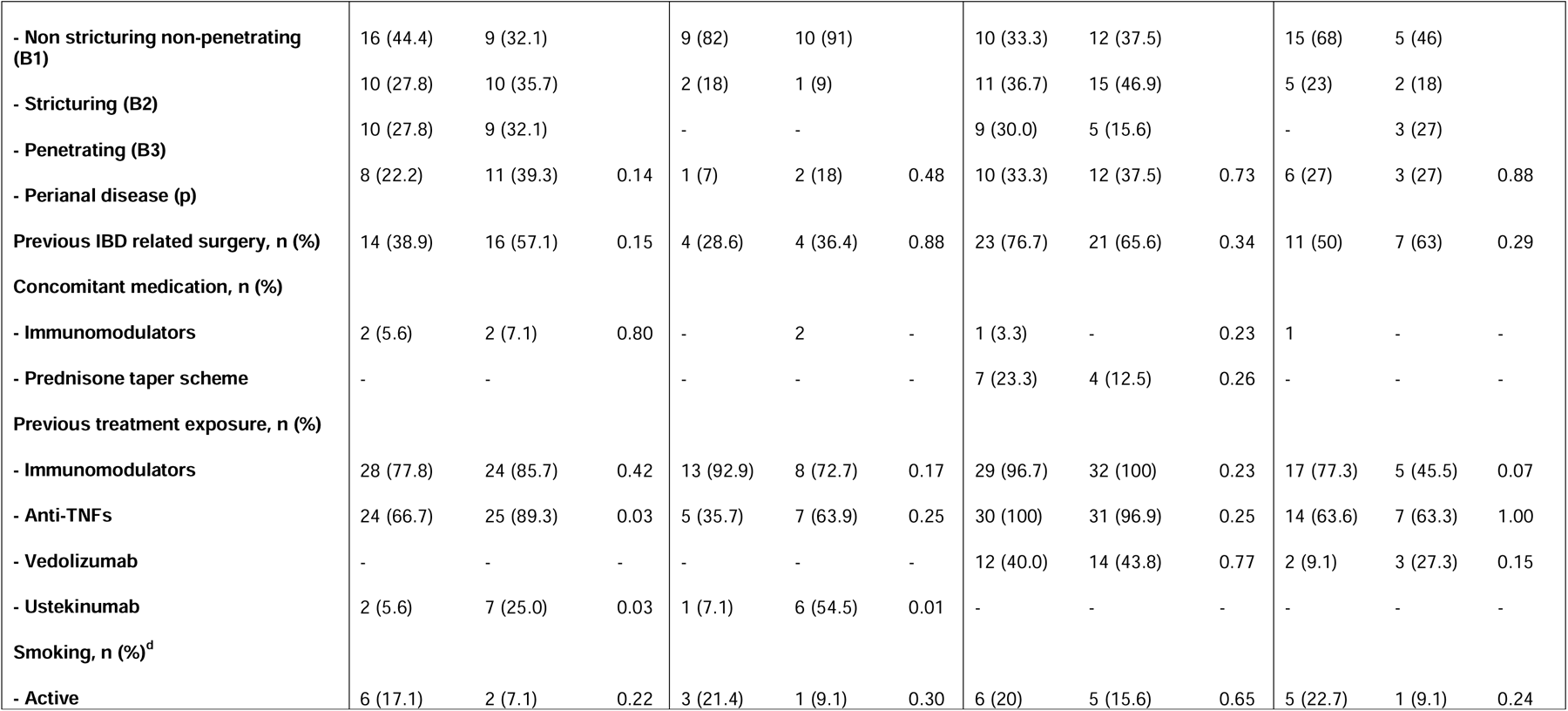
Baseline characteristics discovery- and validation cohorts. Values in bold are significant, percentages shown are valid percentages. ADA: adalimumab. VDZ: vedolizumab. USTE: ustekinumab. R: responder. NR: non-responder, SD: standard deviation. IQR: interquartile range, CRP: C-reactive protein. FCP: Fecal calprotectin. HBI: Harvey Bradshaw Index. SES-CD: simple endoscopic disease activity score, Immunomodulator: azathioprine, mercaptopurine, thioguanine, methotrexate. Anti-TNF: infliximab, adalimumab or golimumab.

### Blood DNA methylation profiling predicts response to vedolizumab and ustekinumab

To identify prognostic biomarkers of VDZ and USTE response, we performed supervised machine learning through stability selected gradient boosting^34,35^ on blood samples obtained shortly before the start of treatment (**Fig. 1a**). The model was trained on the discovery cohort acquired in Amsterdam and subsequently validated in the validation cohort acquired in Oxford. We were able to generate response-predicting models with an area under the curve (AUC) of 0.87 with a standard deviation of 0.06 and 0.89 with a standard deviation of 0.08 for VDZ and USTE, respectively, when testing against the discovery cohort (**Fig. 1b**). The models comprised 25 and 68 differentially methylated CpGs for VDZ and USTE, respectively (**Fig. 1c, Supplementary Fig. 1-2** and **Supplementary Tables 1-2**). Validating our models against the independent validation cohort yielded an AUC of 0.75 for both VDZ and USTE (**Fig. 1b**), indicating reproducible response-associated differences in DNA methylation prior to the start of treatment.

**Fig. 1:**
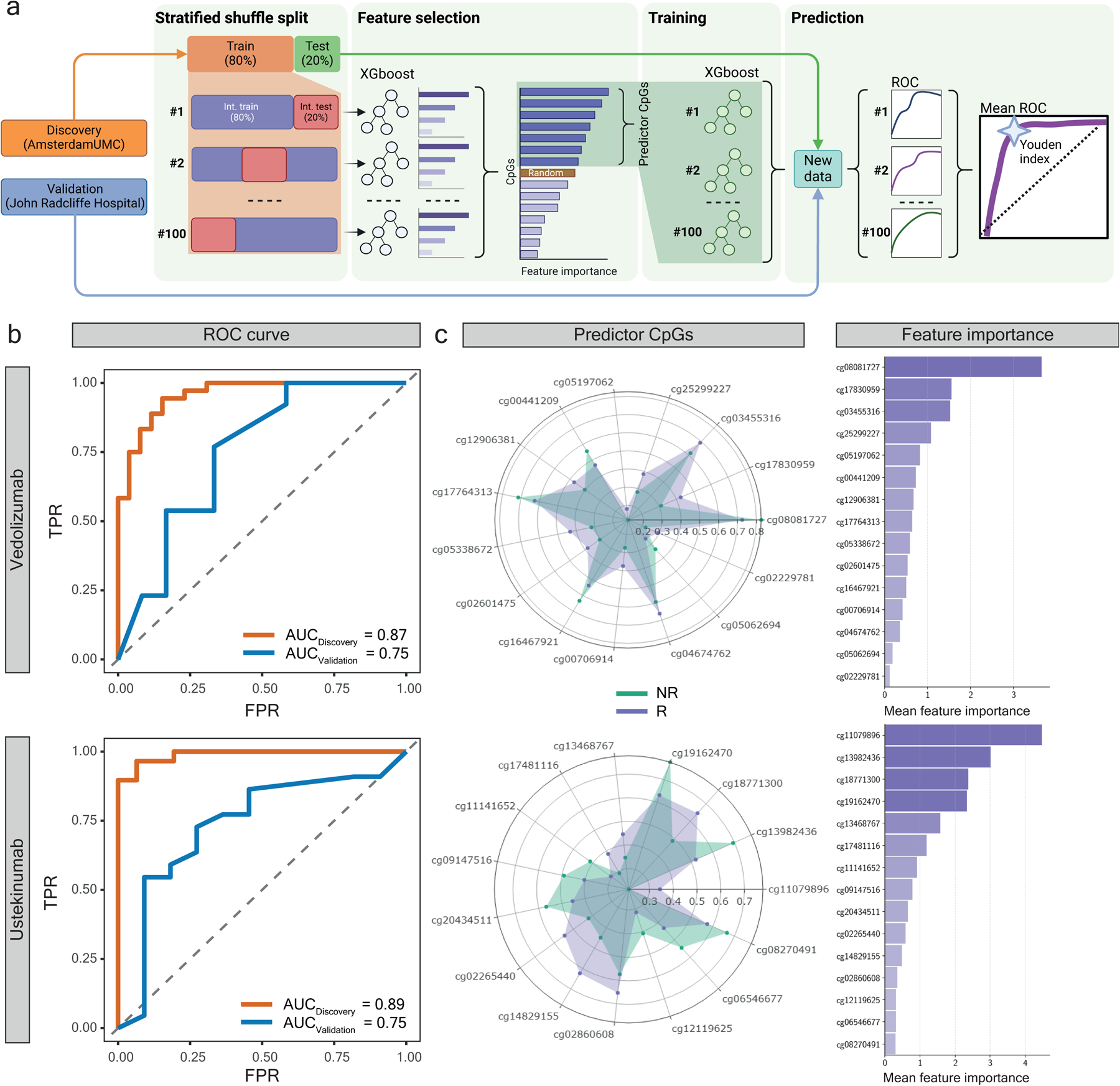
Predictive model using stability selected gradient boosting for response to therapy. **a)** An overview of the feature selection and supervised machine learning approach for predicting response to VDZ and USTE as used in the current study. Int. train: Internal training set. Int. test: Internal test set. ROC: Receiver operator characteristic. **b)** Receiver operating characteristics plots showing the mean area under the curve (AUC) performance of the discovery (n=126) and validation (n=58) cohorts. **c)** Left: radar plots presenting the standardized difference in methylation between R (purple) and NR (green) for the top 15 predictor CpGs. Right: Aggregated feature importance of the top 15 predictor CpGs.

While the discrepancy between discovery and validation performances is expected as part of machine learning, we hypothesized that the differences were partially due to differences in response assessments. During the COVID-19 pandemic, endoscopic assessments were scheduled less frequently resulting in the validation cohort consisting of patient samples in whom response was defined on both strict (combined clinical-, biochemical- and endoscopic evaluation) or modified criteria (combined clinical- and biochemical evaluation). We therefore investigated whether a difference in performance could be observed between both methods of evaluation. This stratification indicated that accuracy was optimized by using the strictly defined combination of clinical- and endoscopic endpoints for both VDZ (AUC_strict_ = 0.83 versus AUC_modified_ = 0.66) and USTE (AUC_strict_ = 0.83 versus AUC_modified_ = 0.72) (**Supplementary Fig. 3a**). Application of the strict criteria yielded performances similar to the discovery dataset thereby confirming that our model is likely more capable at predicting response as defined using combined clinical-, biochemical- and endoscopic evaluations.

In addition, we were interested whether the performance of our models was affected by prior exposure to anti-TNF medication. We observed a better performance of our models among anti-TNF naïve compared to anti-TNF exposed patients for both VDZ (AUC_non-exposed_ = 0.85 versus AUC_exposed_ = 0.66) and USTE (AUC_non-exposed_ = 0.97 versus AUC_exposed_ = 0.63) (**Supplementary Fig. 3b**).

Focusing on the practical implications in a clinical setting, we calculated a sensitivity of 0.769 and a specificity of 0.67 for VDZ and both a sensitivity and specificity of 0.73 for USTE (**Table 2**). Next, we computed the likelihood ratio of response and the post-test probability of response to aid clinicians in accurately predicting response following a positive test outcome. From Lowenberg *et al.* ^7^, we note that 50 of the 110 VDZ-treated CD patients presented with endoscopic response at week 52 indicating a pre-test probability of response of 0.45. At the calculated sensitivity of 0.77 and a specificity of 0.67, the likelihood ratio of response is 2.31, thereby making the post-test probability of response 0.65 for VDZ. Similarly, for USTE, prior research showed that 75 of the 179 USTE-treated CD patients presented with endoscopic response at week 52^8^, indicating a pre-test probability of response of 0.42. At the calculated sensitivity and specificity of 0.73 the likelihood ratio is 2.67 and the post-test probability of response is 0.66. Taken together, the probability that a patient would actually respond to VDZ or USTE when classified as responder is 0.20 and 0.24 higher than the current standard of care.

**Table 2.** Predictive performance metrics of the prognostic biomarkers on the discovery cohort acquired at the AmsterdamUMC (n=126) and the validation 8 cohort acquired at the John Radcliffe Hospital (n=58). TP = True positives. TN = True negatives. FP = False positives. FN = False negatives. AUC = Area under the receiver operator characteristic curve. VDZ: vedolizumab. USTE: ustekinumab.

|  | VDZ<br>discovery | VDZ<br>validation | USTE<br>discovery | USTE<br>validation |
| --- | --- | --- | --- | --- |
| TP | 33 | 10 | 27 | 16 |
| TN | 22 | 8 | 29 | 8 |
| FP | 4 | 4 | 2 | 3 |
| FN | 3 | 3 | 2 | 6 |
| AUC | 0.87 | 0.75 | 0.89 | 0.75 |
| Recall | 0.92 | 0.77 | 0.93 | 0.73 |
| Precision | 0.89 | 0.71 | 0.93 | 0.84 |
| F1-score | 0.90 | 0.74 | 0.93 | 0.78 |

### Methylation status of predictor CpGs remains stable over time

As baseline samples were acquired pre-treatment, we sought to understand whether the initiation of treatment affected the methylation status of the predictor CpGs. To this end, we compared samples obtained at the time of response assessment (T2) with samples obtained pre-treatment (T1). We could not identify any statistically significant differences in DNA methylation levels for any of the predictor CpGs (**Fig. 2a**). Indeed, comparing the differences over time suggested that the mean difference between R and NR was similar both pre-treatment and at response assessment (**Fig. 2b**). Furthermore, a two-way, mixed, consistency intra-class correlation (ICC) analysis indicated highly-stable DNA methylation over time with 24 out of 25 VDZ and 62 out of 68 USTE predictor CpGs presenting ICC values ≥0.75 (**Fig. 2c**, **Supplementary Fig. 1 and 2**). This observation was corroborated by interrogating our previous longitudinal consistency analysis of peripheral blood DNA methylation from 46 adult IBD patients collected at 2 time points with a median of 7 years (range, 2-9 years) in between^36^. Here, we observed that the majority (16 out of 25 VDZ and 52 out of 68 USTE) of the predictor CpGs presented “good” (0.75 ≤ ICC < 0.9) to “excellent” (0.9 ≥ ICC) stability^37^ over a median span of 7 years (**Fig. 2c**). As a final validation, we utilized the prognostic model to predict response to therapy of the samples obtained at response assessment, where we obtained better performances compared to the pre-treatment samples (AUC_VDZ_ = 0.97; AUC_USTE_ = 1.00) (**Fig. 2d**). Altogether, our observations suggest that response-associated differences in DNA methylation detected prior to treatment remain stable during treatment.

**Fig. 2:**
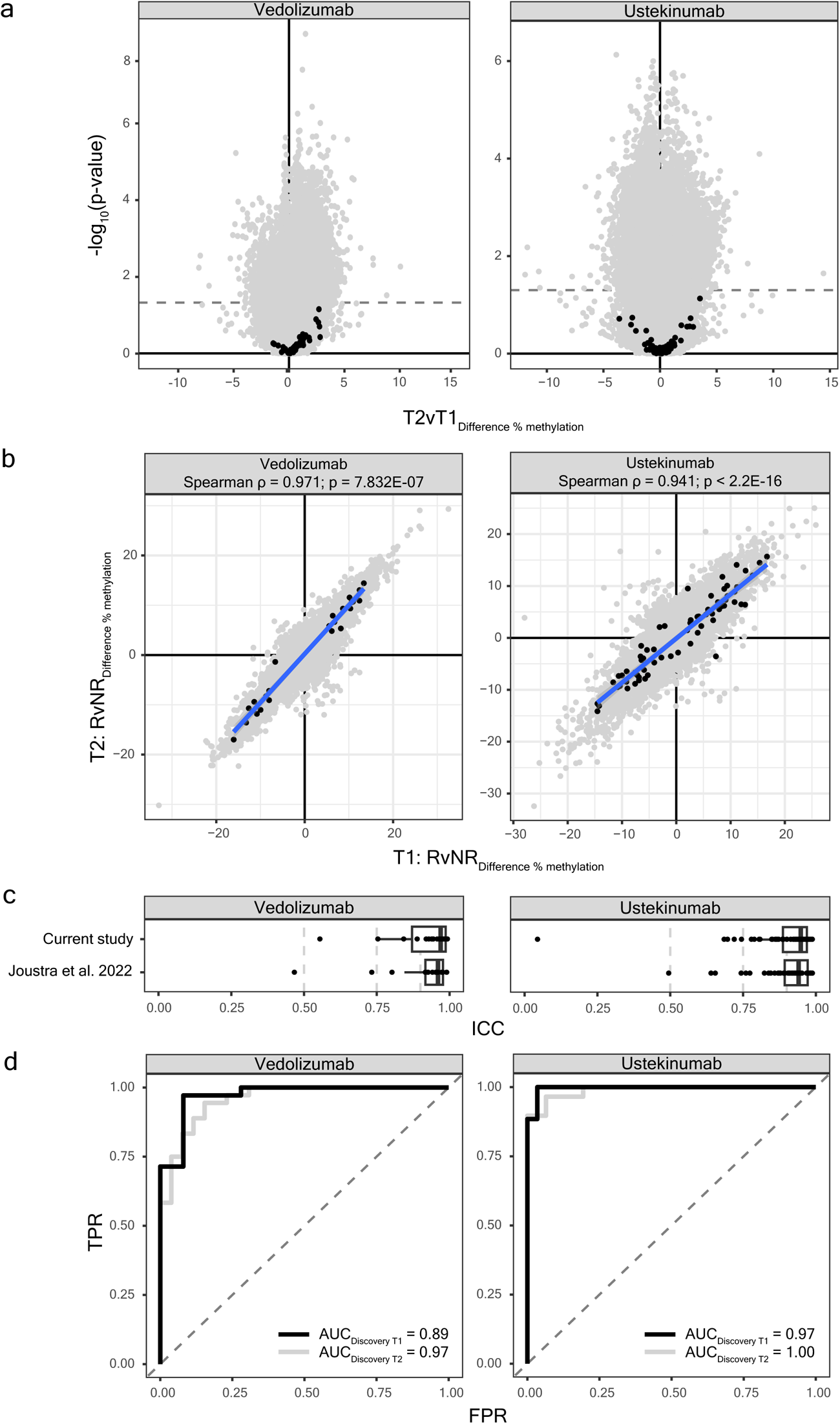
Longitudinal stability analyses. **a)** Volcano plot representing the differential methylation analyses when comparing into treatment (T2) with pretreatment (T1) where grey dots represent CpG loci located on the Illumina HumanMethylation EPIC BeadChip array and black dots represent response-associated predictor CpGs. X-axis represents mean difference in percentage methylation, Y-axis represents the statistical significance as depicted in –log_10_(p-value) with a dashed line at p-value = 0.05. **b)** Scatterplot showing the correlation of differential DNA methylation between R and NR pretreatment (T1) and into treatment (T2). Grey dots represent CpG loci located on the Illumina HumanMethylation EPIC BeadChip array and black dots represent response-associated predictor CpGs. **c)** Boxplot of the two-way, consistency, intra-class correlation (ICC) coefficients of the predictor CpGs calculated when comparing pretreatment and into treatment as well as the ICC coefficients of the predictor CpGs obtained from a previous study on long-term stability of DNA methylation in IBD patients^53^. The vertical dashed grey lines represent classification boundaries introduced by Koo and Li^37^, with blocks representing poor (ICC < 0.5), moderate (0.5 ≤ ICC < 0.75), good (0.75 ≤ ICC < 0.9), and excellent (0.9 ≥ ICC). **d)** Receiver operating characteristic plots representing the predictive performance into treatment (T2; black) and pre-treatment (T1; grey) as reference.

### Patients who previously experienced treatment failure to multiple biological treatments are classified correctly

Having established that the prognostic response prediction models for both drugs perform well both pre- and into treatment, we explored the performance of our model in a further independent cohort of 33 adult CD patients from whom blood had been collected after sequential treatment with anti-TNF, VDZ, and/or USTE. Treatment failure was determined with endoscopy in 28 (82%), MRI in 2 (6%), or the need for surgical resection in 4 (12%), combined with biochemical biomarkers such as CRP in 12 (35%) and/or FCP in 16 (47%) and clinical parameters in 32 (94%) patients. In this cohort, a relatively large proportion of patients had extensive disease location (67.6%), perianal disease (52.9%), and/or IBD-related surgery (70.6%) in the past (**Supplementary Table 3**).

Both VDZ and USTE response prediction models accurately predicted NR, with 24 out of 27 (88.9%) VDZ-NR patients and 24 out of 26 (92.3%) USTE-NR patients correctly identified. Focusing specifically on the patients with previous non-response to both VDZ and USTE the models correctly classified 16 out of 19 (84%) as NR. Notably, 12 of out of these 19 patients (75%) were anti-TNF experienced.

### Assessment of potential confounding variables

Through multiple linear regression analyses we observed that 22 of the 25 (88%) VDZ response-associated CpGs and 38 of the 68 (55%) USTE response-associated CpGs (**Fig. 3a** and **Supplementary Fig. 1-2**) presented statistically significant differences. The large discrepancy between the linear regression analyses and the USTE response-associated CpGs indicates that a more complex non-linear relationship exists among the response-associated predictor CpGs and underscores that statistical p-values may not equal biologic or functional relevance and/or importance at an individual level. As it has been established that the peripheral blood DNA methylome is associated with certain phenotypic characteristics such as sex, age, smoking status, as well as the underlying cellular composition^38–41^, we investigated whether any of these variables confounded our results. In doing so with linear regression analysis, we observed that 12 out of 22 (55%) and 30 out of 38 (79%) markers remained significantly associated with response for VDZ and USTE, respectively (**Fig. 3a**). In terms of effect size, the mean percentage methylation difference between R and NR of the predictor CpGs on average decreased by 15% and increased by 5% in VDZ and USTE, respectively (**Fig. 3b**). To understand whether the confounding variables are capable of predicting response, we constructed a prediction model solely based on the confounding variables using the discovery cohort and tested this on the validation cohort. The confounder model yielded an AUC of 0.57 and 0.65 for VDZ and USTE, respectively, against the validation cohort indicating worse performance than the prediction model based on CpGs only (**Fig. 3c**). Our results indicate that while there appears to be a significant association with sex, age, smoker status and blood cell distribution, the CpG-only prediction model outperforms a confounder-only model.

**Fig. 3:**
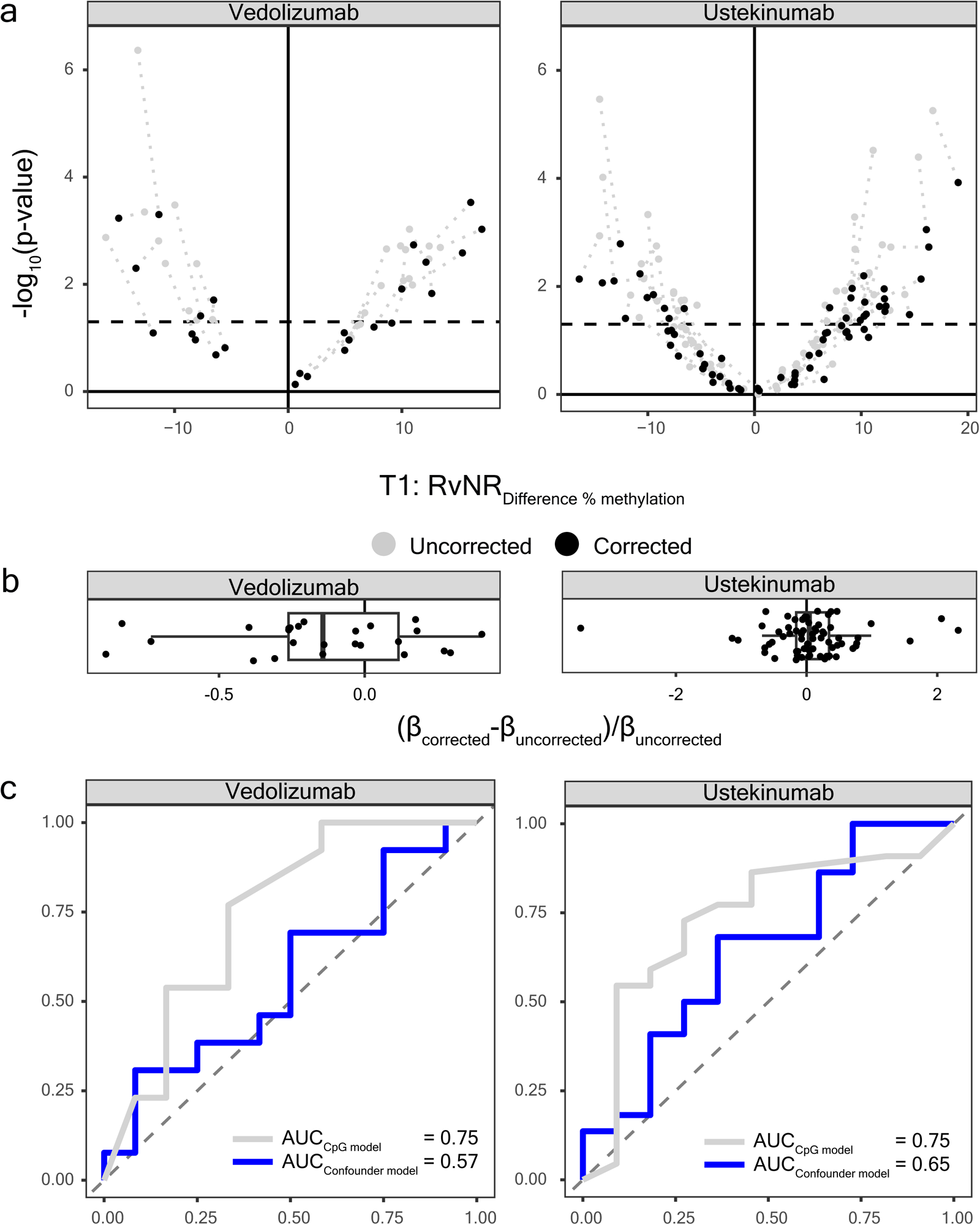
Analyses of potential confounding variables. **a)** Volcano plot representing the change in response-associated differential methylation when correcting for the potential confounding variables age, sex, and estimated cellular composition (black) or not (grey). X-axis represents mean difference in percentage methylation, Y-axis represents the statistical significance as depicted in –log_10_(p-value). The dotted line represents a threshold set at p = 0.05. **b)** Boxplot of the change in effect size after correcting for the confounding variables as calculated by (β_corrected_-β_uncorrected_)/ β_uncorrected_. **c)** Receiver operator characteristic curve comparing the response-prediction model for the CpG model (grey) with the confounder model (blue).

We next investigated whether the predictor CpGs were significantly associated with severity of systemic and intestinal inflammation at baseline measured using CRP and FCP, as was previously reported by Somineni *et al.*^42^. For VDZ, 5 predictor CpGs significantly associated with CRP whereas only a single CpG was associated with FCP (**Supplementary Fig. 4**). For USTE, we observed 9 predictor CpGs associated with CRP and 2 predictor CpGs with FCP (**Supplementary Fig. 4**). In all cases, mean differences in methylation were smaller than 0.5%. These observations suggest that the majority of the identified predictor CpGs are independent of inflammatory status at onset of therapy.

### RFPL2 presents concordant response-associated differential methylation and expression

To understand the functional relevance of the predictor CpGs, we annotated the them to their respective genes based on whether they were located in either a gene promoter or enhancer resulting in the annotation of 20 VDZ response-associated predictor CpGs to 16 unique genes (**Supplementary Table 1**) and 43 USTE response-associated predictor CpGs to 46 genes (**Supplementary Table 2**). We performed transcriptomic analyses on a subset of samples from the discovery cohort (VDZ; N_T1_=10_R_10_NR_, N_T2_=10_R_8_NR_ and USTE; N_T1_=14_R_11_NR_, N_T2_=10_R_9_NR_). Comparing R with NR identified pretreatment differences for predictor CpG-associated genes *TULP4* (p-value_T1_ = 4.38E-02) and *RFPL2* (p-value_T1_ = 4.72E-02) for VDZ (**Fig. 4a, b** and **Supplementary Table 4**), with *RFPL2* presenting a significant inverse correlation between DNA methylation and gene expression (Pearson r = -0.65; p-value = 1.39E-03) (**Fig. 4c**). For USTE, we observed significant differences in the expression of predictor CpG-associated genes *MRC1* (p-value_T1_ = 4.25E-04) and *TMEM191B* (p-value_T1_ = 7.31E-03) (**Fig. 4a** and **d** and **Supplementary Table 5**) with *TMEM191B* presenting a significant positive correlation between DNA methylation and gene expression (Pearson r = 0.57; p-value = 3.11E-03) (**Fig. 4e**). We next investigated whether predictor CpG-associated genes presented differential expression after the start of treatment by comparing R with NR at response assessment. VDZ predictor CpG-associated genes *MCM2* (p-value_T2_ = 2.40E-03) and *RFPL2* (p-value_T2_ = 3.88E-03) were differentially expressed at response assessment (**Fig. 4a, f** and **Supplementary Table 4**) with *RFPL2*, once again, presenting a significant inverse correlation between DNA methylation and expression (Pearson r = -0.55; p-value = 0.017) (**Fig. 4g**). For USTE, predictor CpG-associated genes *POTEF* (p-value_T2_ = 1.47E-02), *HDAC4* (p-value_T2_ = 2.18E-02), *PARP4* (p-value_T2_ = 3.49E-02) and *MARK3* (p-value_T2_ = 2.87E-02) presented differential expression at response assessment (**Fig. 4a, h** and **Supplementary Table 5**), but did not show any significant correlation with DNA methylation. Taken together, our results show that some of the response predictor CpG-associated genes present differential expression either pretreatment and/or during response assessment, with VDZ- and USTE-response-associated genes *RFPL2* and *TMEM191B*, respectively, presenting a significant correlation between DNA methylation and gene expression. Nonetheless, we acknowledge that most predictor CpG-associated genes present no response-associated differential expression.

**Fig. 4:**
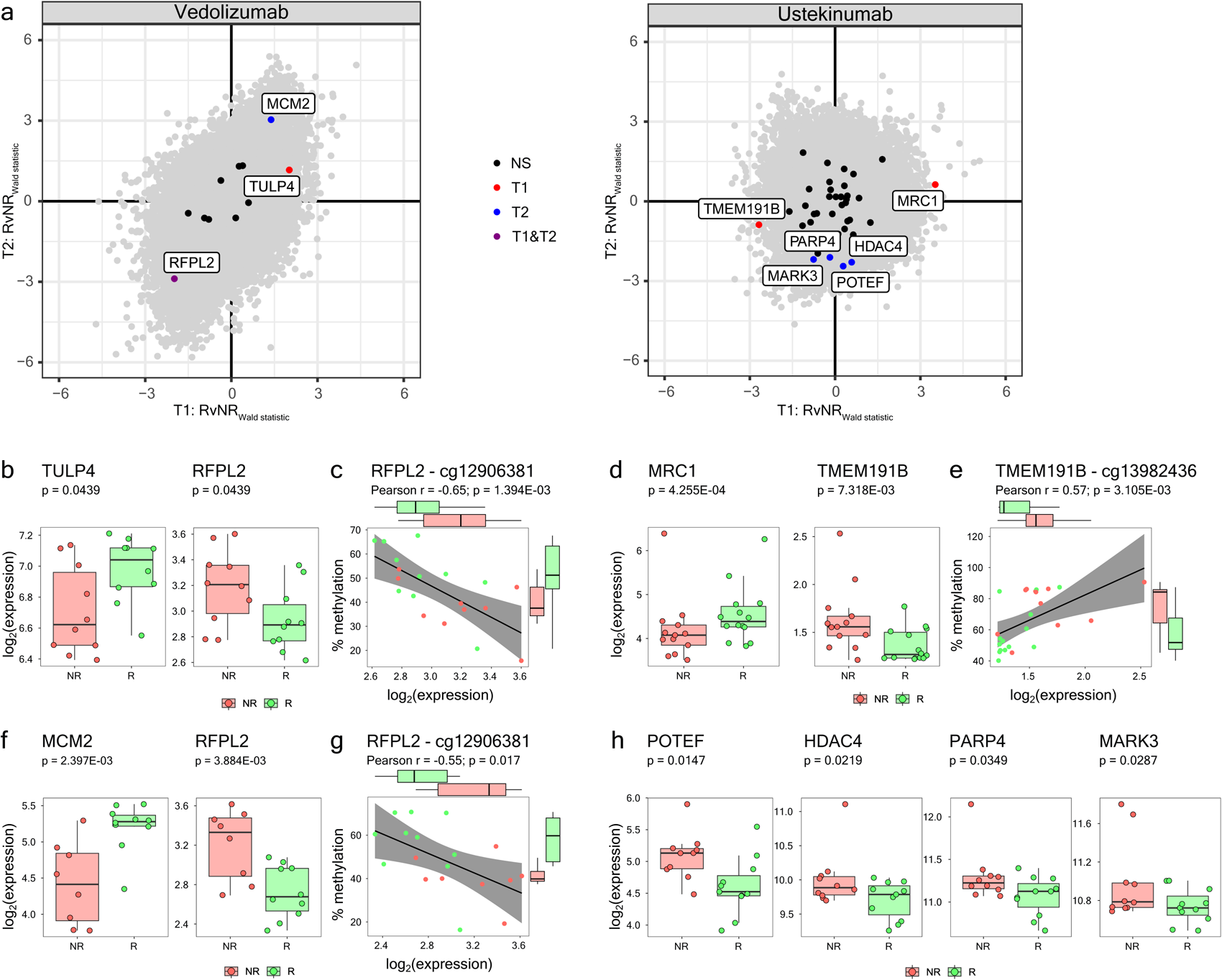
Integrative analyses of predictor CpG-associated genes. **a)** Scatterplot showing the effect size (Wald statistic) of the response-associated difference in pretreatment (T1; X-axis) and into treatment (T2; Y-axis). Grey dots represent all genes measured, black dots represent non-differentially expressed predictor CpG-associated genes, red, blue and purple dots represent predictor associated genes that are differentially expressed at T1, T2, or T1 and T2, respectively. **b)** Boxplot of the T1 log_2_(expression) for VDZ response predictor CpG-associated genes *TULP4* and *RFPL2* stratified by response. **c)** Scatterplot of the T1 log_2_(expression) of *RFPL2* (X-axis) relative to the percentage DNA methylation (Y-axis) for the predictor CpG cg12906381 annotated with the Pearson correlation coefficient and the associated p-value. **d)** Boxplot of the T1 log_2_(expression) for USTE response predictor CpG-associated genes *MRC1* and *TMEM191B* stratified by response. **e)** Scatterplot of the T1 log_2_(expression) of *TMEM191B* (X-axis) relative to the percentage DNA methylation (Y-axis) for the predictor CpG cg13982436 annotated with the Pearson correlation coefficient and the associated p-value. **f)** Boxplot of the T2 log_2_(expression) for VDZ response predictor CpG-associated genes *MCM2* and *RFPL2* stratified by response. Scatterplot of the T2 log_2_(expression) of *RFPL2* (X-axis) relative to the percentage DNA methylation (Y-axis) for the predictor CpG cg12906381 annotated with the Pearson correlation coefficient and the associated p-value. g) Boxplot of the T2 log_2_(expression) for USTE response predictor CpG-associated genes *POTEF, HDAC4, PARP4* and *MARK3* stratified by response.

## Discussion

Although the introduction of biologicals has transformed the care of patients with CD, the current clinical practice of treatment selection remains suboptimal. Nonetheless, a large body of real-world evidence studies suggests there could be a place for a more individualized approach^43,44^. Therefore, in parallel with ongoing drug development, predictive biomarkers that allow for selection of successful medical therapy would represent a major step forward in clinical care.

Here, we conducted a longitudinal case-control study where we identified methylation signatures composed of 25 and 68 markers associated with combined endoscopic, biochemical and clinical response to VDZ and USTE, respectively, in a cohort of adult CD patients in whom treatment response was assessed with stringent criteria. We were able to build models with significant predictive performance at an AUC > 0.85 for both models. The models demonstrated similar performance in an independent, external validation with an AUC of 0.75 for both models.

Recent real-world data and (post-hoc) findings from both the GEMINI and UNITI trials indicate superior response to both VDZ and USTE in anti-TNF naïve patients^45–49^. In our discovery cohorts, 77% of VDZ and 98% of the USTE-treated patients were previously exposed to anti-TNF medication. Stratifying the patients in the validation cohort by previous anti-TNF exposure showed that both models performed noticeably better in anti-TNF naïve rather than exposed patients. However, the number of patients included in both subset comparisons are relatively small and further exploration using larger groups patients are needed. Furthermore, we demonstrate the ability of our models to effectively identify patients with previous non-response to both VDZ and USTE treatment. This holds true for both anti-TNF naïve and experienced patients, providing significant importance for clinical practice, as the precise prediction of non-response to both drugs offers clinicians the opportunity to make informed decisions. For anti-TNF experienced patients, this could mean a direct switch towards newer modes-of-action (i.e. JAK-inhibitors).

While this study is the first to demonstrate the utility of DNA methylation profiling in whole blood for objective response to VDZ and USTE in CD patients, previous analyses from two separate studies explored its application to predict anti-TNF response in both CD and UC patients^50,51^. In the first study, the authors sought to identify an anti-TNF response-associated profile combining integrated methylation and gene expression data from samples taken before and 2 weeks into treatment using a primary endpoint of clinical remission at week 14^50^. The observations were made using a mixed cohort of 37 IBD (18 CD, 19 UC) patients and validated against a publicly available gene expression data of 20 CD patients. In the second study, the authors did not show replication of these observations using methylation data of 385 patients, as part of the previously published PANTS study^51,52^. In this study, bio-naïve patients with active CD started treatment with adalimumab or infliximab. Interestingly, the authors report 323 differentially methylated positions annotated to 210 genes identified at baseline that were significantly associated with serum drug concentrations at week 14. This profile could potentially be of interest to identify patients in need of intensified anti-TNF drug monitoring or dosing. The methodological differences and lack of endoscopic data in the previous experiments preclude direct comparison with our study in which outcome assessments were more stringent.

Through two separate stability analyses, we demonstrated both short- and long-term hyper stability of the majority of our identified CpG markers indicating their independence of treatment as well as the resultant difference in inflammation. The latter is further evidenced by the lack of correlation between the methylation status of the predictor CpGs and both baseline CRP and FCP, therapy switch or even CD-related surgery^53^ suggesting that the CpGs are response-predictors but do not directly mediate inflammation. Nonetheless, confounder analysis indicated potential confounding for some of the predictor CpGs for age, sex, smoking status and the blood cell distribution. We observed however that prediction modeling using the confounders only performed significantly worse than the CpG model, indicating that confounders do not contribute substantially to the predictive performance.

We note that the predictor CpGs annotate to genes associated with MHC class I (*HLA-C*) and cell migration (*TSPEAR, NID2*)^54,55^ for VDZ and macrophage function (*RHOJ*, *MARK3* and *PCGF3*)^56–61^ and polarization (*PKNOX1* and *MRC1*)^62–64^, histone remodeling and Th17-differentiation (*HDAC4*)^65–69^ and TGF-β signaling (*SMAD1* and *EBF3*)^70–77^ for USTE. Integrating DNA methylation with their transcriptomic data indicates that VDZ-response-associated *RFPL2*, which encodes an E3 ubiquitin ligase^78,79^, and USTE-response-associated *TMEM191B*, which encodes a transmembrane protein, present concordant differential methylation and expression. However, their exact role in either IBD and/or response to therapy remains unknown to date. Accordingly, beyond the utility of the predictor CpGs in classifying response to therapy, identifying their role in the pathogenesis and etiology of non-response remains challenging at present and hence a subject for future studies.

Our study derives its main strength from the sampling and endpoint assessment strategy. In addition, patients with anti-drug antibodies or without a measurable serum drug concentration and those that stopped treatment due to adverse events were excluded prior to the selection of this cohort. Non-responders therefore reflect a more homogenous group of true biological (i.e. pharmacodynamic) non-response rather than failure due to pharmacokinetics or intolerance. Second, the relatively small sample size was justified by performing a sample size estimation based on prior pilot experiments in combination with previous studies on statistical power in EWAS^80^. Lastly, besides stability of the observed methylation differences during induction- and maintenance treatment, our markers demonstrated stable differences between R and NR over time, which was further corroborated when interrogating our 7-year longitudinal DNA methylation survey. This time-independent behavior of the predictor CpGs indicates that exposure over time and change in inflammatory parameters does not affect the response-associated behavior of the CpGs, suggesting that the CpGs are very stable, which in turn increases its utility in clinical practice.

There are however some limitations to this study. First, a post-hoc power analysis of the mean effect size difference between responders and non-responders indicated a mean absolute difference of 9.9% and 7.6% for VDZ and USTE, respectively. At the collected samples sizes, this would translate to a statistical power of approximately 98.4% and 64.3% for VDZ and USTE, respectively^80^. Despite the lower power for USTE, we note that the model remained performant in predicting response in the external cohort. Second, in the external cohort response assessment was less stringent in approximately 70% of the patients due to the COVID-19 pandemic during which we encountered a notable reduction in the access to non-essential endoscopies, particularly in the UK^81^. Nonetheless, the modified response criteria are a reflection of clinical parameters that physicians commonly use in daily practice. This pragmatic approach enhances the overall generalizability of our results to a much larger IBD population. Notably, the analysis where we included the available endoscopic outcomes in the subset of UK patients for whom these data were available, enhanced the performance to an AUC of 0.83 for both the VDZ and USTE models, reinforcing the validity of both models in identifying objective responders to these biological therapies. While we acknowledge this limitation, we believe that our comprehensive approach provides valuable insights into the predictive capabilities of the models under diverse clinical scenarios. Third, while we purposely used PBL samples as these are minimally invasive and easily obtained during daily clinical practice, PBL represents a mixed cellular population. Therefore, the specific cell types responsible for the observed predictive signal remain unidentified^82^. It should be noted however that after correcting for the blood cell distribution, several predictor CpGs remained statistically significant, indicating independence of cellular composition. Third, the majority of the predictor CpG loci identified are situated within gene introns, complicating the biological interpretation of our findings. Although the identified predictor CpGs collectively serve as a strong predictor of response, we acknowledge that the underlying biology behind this observation is more complicated than merely the inverse correlation between DNA methylation and gene expression. Lastly, while strict removal of most catalogued and predicted genetic variant-binding probes, we acknowledge that there is still a possibility that underlying genetic differences could have influenced our outcome^83^. Nonetheless both models performed effectively in both the discovery-(Dutch) and the validation (UK) cohorts.

In summary, our findings pave the way towards personalized medicine for CD. Several US and European studies report a significant reduction in pharmaco-economic burden of CD if clinical and endoscopic remission can be achieved with adequate treatment compared to the cost of treatment of IBD patients on suboptimal medication^84–88^. In the absence of a predictive biomarker panel, current endoscopic response rates at week 52 have been reported around 45% in VDZ and 42% in USTE treated CD patients^7,8^. Taken together, our biomarker panels could potentially increase these proportions by approximately 20% for VDZ and 24% for USTE, thereby significantly impacting healthcare costs and disease burden in these patients. We acknowledge that clinical validation of our findings in a randomized prospective trial, comparing our method of pre-treatment selection with current clinical practice, is needed to firmly demonstrate both clinical and economic benefit^89^. To this end, the Omicrohn trial as part of the ongoing Horizon Europe funded METHYLOMIC project has been launched and is currently underway^90^.

## Online Methods

### Study population and design

We prospectively recruited adult male and female CD patients that presented with a combination of clinical, biochemical and endoscopic disease activity at ileo-colonoscopy and were scheduled to start VDZ or USTE treatment within 1 year after the last endoscopy at the Amsterdam University Medical Centers, University of Amsterdam, Amsterdam, Netherlands (discovery cohort) and the John Radcliffe Hospital, Oxford, United Kingdom (validation cohort). All patients were naïve to the biological of interest.

Patients were treated according to standard-of-care protocols, which for VDZ meant that patients were given 300 mg infusions at week 0, 2 and 6 followed by infusions at an 8 week interval. For USTE, standard-of-care involved patients receiving a single intravenous infusion (6 mg/kg rounded to 260 mg, 390 mg or 520 mg) at week 0 and subsequent 90 mg subcutaneous injections at an 8 week interval. For both VDZ and USTE, interval intensification every 6 or 4 weeks with an additional week 10 infusion for VDZ or extra intravenous boost infusion for USTE at the treating physicians’ discretion. To ensure assessment of mechanistic and not pharmacokinetic failures of each biological treatment, only patients with measurable serum concentrations without anti-drug antibodies at response assessment were used for methylation analyses.

The project was approved by the medical ethics committee of the Academic Medical Hospital (METC NL57944.018.16 and NL53989.018.15) and written informed consent was obtained from all subjects prior to sampling, as well as by the National Health Service Research Ethics committee. (REC reference: 21/PR/0010 Protocol number: 14833 IRAS project ID: 266041). UK patients were recruited and consented under the ethics of the Translational Gastrointestinal Unit biobank IBD cohort ethics (09/H1204/30) and GI cohort ethics (16/YH/0247 and 21/YH/0206). The quality of the collected data and study procedures were assessed by an independent monitor.

### Vedolizumab discovery and validation cohort characteristics

The VDZ discovery cohort consisted of 64 patients (N_R_ = 36, N_NR_ = 28) of which 49 (77%) had previously been exposed to anti-TNF treatment and 9 (14%) to USTE. Response was defined on endoscopic as well as clinical/biochemical criteria in this group (see section “Definitions of response”). In 62 (97%) patients, a follow-up endoscopy was performed. The remaining 2 patients were classified as non-responders as a result of urgent surgical intervention due to worsening of disease without CD-associated complications, such as stenosis or perforating disease. R and NR presented overall comparable clinical characteristics, with no significant differences in age, sex and smoking behavior. Importantly, serum VDZ concentrations at T2 were not significantly different between R and NR (median 15 (IQR 7.7-20.5) versus median 14 (IQR 3.6-27.5), p-value = 0.77) although more NR patients received an additional VDZ infusion at week 10 (35.7% vs 13.9%, p-value = 0.04). Notably, patients in the NR group had more frequently been exposed to anti-TNF treatment (89.3% vs 66.7%, p-value = 0.03) and/or USTE (25% vs 5.6%, p-value = 0.03) compared to R group. Thirteen of the 15 R patients received VDZ as first-line biologic, had a shorter disease duration, lower rates of previous surgery and perianal disease as well as a higher percentage of B1 phenotype compared to anti-TNF experienced patients.

The VDZ validation cohort consisted of 25 additional patients (N_R_ = 14 and N_NR_ = 11) from Oxford with a median 9 (IQR 3-15) year disease duration. Seventeen (68%) did not undergo a follow-up colonoscopy to assess endoscopic response due to restrictions during the COVID-19 pandemic and therefore were assessed using the modified definition of response, which was defined as a combination of clinical and biochemical parameters (see “Definitions of response” below). Out of these 25 patients, 13 (52%) were biological naïve, 12 (48%) were anti-TNF-experienced and 7 (28%) were previously treated with USTE, which was enriched among the NR group (54.5% vs 7.1%, p-value = 0.01). All other clinical characteristics were comparable in R and NR, including age, sex, and smoking behavior. As in the VDZ discovery cohort, the majority (69%) of biological naïve patients were responders to VDZ.

### Ustekinumab discovery and validation cohort characteristics

The USTE discovery cohort consisted of 62 patients (N_R_=30, N_NR_=32) of which 60 (98%) were previously exposed to anti-TNF and 26 (42%) to VDZ. A follow-up endoscopy was performed in 58 (94%) patients. The remaining 4 were evaluated with serial intestinal ultrasound using validated criteria assessed by an expert IBD ultrasonographist. Clinical characteristics did not significantly differ between R and NR, although the R population consisted of more female patients (R = 80%, NR = 56.3%, p-value = 0.05). No significant differences in treatment intensification (21.9% vs 6.7%, p-value = 0.08), extra intravenous boost infusions (p-value = 0.26) or serum USTE concentrations at T2 between R and NR were observed (median 3.0 (IQR 1.8-5.5) versus median 4.8 (IQR 2.1-8.6), p-value = 0.31).

The USTE validation cohort consisted of 33 patients (N_R_=22 and N_NR_=11), with a median disease duration of 11 (IQR 3-20) years. Twenty-five (76%) did not undergo follow-up colonoscopy as a result of pandemic-related restrictions on non-essential endoscopy and therefore were assessed using the modified definition of response. Of the 33 included patients, 9 were biological naïve (52%), 21 (63.6%) were anti-TNF-experienced and 5 (15.2%) were previously treated with VDZ. Responders presented a significantly longer disease duration compared to non-responders (median 15 vs 5 years, p=0.05). No significant differences in age, sex, and smoking behavior were observed.

### Sample collection and storage protocols

In all patients, whole peripheral blood leukocyte (PBL) samples were collected for measurement of epigenome-wide DNA methylation prior to the start of VDZ/USTE, before the baseline endoscopy or the first infusion (time point 1) and after an interval of 6-9 months into treatment (time point 2) using 4.0-6.0mL BD ethylenediaminetetraacetic acid (EDTA) vacutainer tubes. For the Amsterdam discovery cohort, samples were subsequently aliquoted into 1.10mL micronic tubes before storing at -80 °C until further handling. The Oxford validation samples were directly frozen at -80 °C in preparation for later extraction. At both time points, additional PBL samples were stored for the purpose of targeted gene expression analyses from a subset of the Amsterdam discovery patients using 2.0-mL PAXgene Blood RNA tubes and were frozen at –20 °C for 24 h before storing at –80 °C until further handling.

### Definitions of response

At response assessment, patients were classified as responders (R) or non-responders (NR) based on a strict combination of endoscopic, biochemical and clinical criteria: ≥50% reduction in the endoscopic SES-CD score, corticosteroid-free clinical remission (≥3 point drop^91^ in HBI or HBI ≤4 and no systemic steroids) and/or biochemical response (CRP reduction ≥50% or CRP≤5 mg/L and FCP reduction ≥50% or FCP ≤250 µg/g). Modified response was defined as a combination of corticosteroid-free clinical-(HBI ≤4) and biochemical (CRP ≤5 mg/L and/or FCP ≤250 µg/g) remission between week 26-52 without treatment change through week 52.

### DNA isolation and in vitro DNA methylation analysis

For the Amsterdam discovery cohorts, genomic DNA was extracted using the QIAsymphony. We next assessed the quantity of DNA using the FLUOstar OMEGA and quality of the high-molecular weight DNA on a 0.8% agarose gel. Following these steps, 750ng of DNA per sample was randomized per plate, to limit batch effects, after which genomic DNA was bisulfite converted using the Zymo EZ DNA Methylation kit and analyzed on the Illumina HumanMethylation EPIC BeadChip array. Aforementioned work was performed at the Core Facility Genomics, Amsterdam UMC, Amsterdam, the Netherlands.

For the Oxford validation cohorts, genomic DNA was extracted using the Qiagen Puregene Blood Core Kit C at the Oxford Translational Gastroenterology Unit, University of Oxford. DNA samples were assessed for quality using the NanoDrop spectrometer (NanoDrop 1000, Thermo Scientific). A total of 750ng of DNA per sample was randomized per plate, to limit batch effects, after which genomic DNA was bisulfite converted using the Zymo EZ DNA Methylation kit and analyzed on the Illumina HumanMethylation EPIC BeadChip array at UCL Genomics, University College London, London, United Kingdom.

### Raw methylation data pre-processing

DNA methylation data processing was orchestrated in Snakemake (v7.14.1)^92^. Raw methylation data was imported into the R statistical environment (v4.3.1) using the Bioconductor minfi^93,94^ package (v1.44). Quality control was performed using shinyMethyl (v1.38.0)^95^ for probe level quality control, ewastools (v1.7.2)^96^ to ensure paired samples were correctly labeled, and the Horvath clock^97^ to ensure a proper match with the metadata. One patient sample from the VDZ and one patient sample from the USTE discovery cohort were removed due to a discrepancy in the predicted sex with the annotated sex. Raw signals were normalized using functional normalization^98^. Probes were annotated to their gene of interest using the provided Illumina annotations, which were further enhanced with enhancer data as obtained using the publicly accessible promoter-capture Hi-C data^99^. We subsequently calculated the methylation signal in the form of percentage methylation. Technical artefacts as a result of batch, plate and plate position were removed using ComBat (v0.9.7)^100^ as implemented in the sva (v3.50.0)^101^ package, a tool for removing known batch effects in microarray data using the parametric empirical Bayes framework, using the default parameters. Probes hybridizing to allosomes were removed to identify sex-independent differences. Moreover, probes hybridizing to known and tentative genetic variants were removed to identify true methylation signals. Known genetic variants were identified based on their presence in dbSNP (v137), whereas tentative genetic variants were based on the methylation signal displaying a tri- or bimodal distribution, a hallmark of genetic variants underlying DNA methylation^102^, as determined using gaphunter^103^ set to a threshold of 0.25. The eventual number of CpGs used for training the prediction models were 806,308 and 808,815 for VDZ and USTE, respectively. For the validation of the prediction models, raw methylation data from the validation cohort was pre-processed together with the discovery cohort using functional normalization and ComBat to mitigate batch effects introduced by the different experimental setup. From the combined dataset, the predictor CpGs were extracted and the prediction model was recalibrated against the discovery dataset where after predictions were made on the validation dataset.

### Machine learning models: stability selected gradient boosting analyses

The machine learning modeling was divided into two steps, namely feature selection and validation (**Fig. 1a**). Feature selection was performed on the discovery cohort, collected at the AmsterdamUMC, whereas validation was performed on the validation cohort, collected at the John Radcliffe Hospital. During the feature selection procedure, the model was at no point exposed to the validation cohort. To identify baseline epigenetic markers associated with response/non-response to treatment, we implemented a rigorous supervised machine learning approach using stability selected gradient boosting^34,35,104^ combined with covered information disentanglement (CID)^105^. Gradient boosting is an algorithm for supervised learning, which operates through stepwise improvement of weak learners thereby minimizing the overall prediction error against the observed data. We opted for gradient boosting as it captures linear, non-linear and interaction effects better than traditional linear regression approaches^104^. CID, in conjunction with stability selected gradient boosting, represents an approach for feature selection that assigns permutation-based feature importance that, unlike other methods for assigning feature importance, is unbiased by multicollinearity^105^.

We first removed CpGs with low variance and then applied univariate feature selection using a F-test. Stability selection was subsequently employed to identify reliable biomarkers by splitting randomly the discovery data into an 80% training data and a 20% test data using stratified shuffle split and repeating this process 100 times to mitigate overfitting^106^. During each split, we computed the CID for each CpG by randomly permuting it 100 times and calculating the mean feature importance and assessing the average effect of permutation on the model’s performance (i.e., predicted versus true outcome)^105^. After all 100 iterations, the mean feature importance per iteration was averaged and compared against a randomly generated noise variable that was included throughout the entire modeling process where CpGs with an aggregated feature importance ranked above a random variable were termed predictor CpGs and retained for future analyses.

Having determined the predictor CpGs, we subsequently validated the predictive performance internally and externally. Internal validation was performed by extracting the predictor CpGs, training an ensemble of 100 gradient boost models on the 80% discovery training data and utilizing all models to predict against the withheld 20% discovery test data. External validation was performed using a similar approach where the discovery and validation cohort were merged, the predictor CpGs extracted, and an ensemble of 100 models trained on the discovery cohort. The resultant models were then requested to predict the response in the external validation cohort. In both cases, the output of each model yielded a prediction score on a scale of 0 to 1 per sample. The prediction scores were aggregated by calculating the mean prediction score per sample, representing the final, ensembled, output of the model. This final prediction score was used to calculate the receiver operator characteristic (ROC) to assess performance. The resultant prediction scores were subsequently converted into classes by freezing the model at the Youden index, thereby balancing the true positive rate relative to the false positive rate. Analyses were not conducted separately for males and females as the cohorts would become too small. Aforementioned analyses were performed in Python (v3.10) (https://www.python.org), with packages scikit-learn and xgboost for the model development.

### In silico DNA methylation analysis

Differential methylation analyses were performed in R using limma^107^ (v3.46) and eBayes^108^. Separate analyses were run either regressing against the slide and slide position only, or in combination with common confounding variables: age, sex, smoking behavior and the estimated blood cell distribution^109,110^. The blood cell distribution was estimated using the method described by Houseman *et al.* ^109^ and implemented by Salas *et al*.^110^ for the Illumina HumanMethylation EPIC BeadChip array, where methylation profiles of the current data are compared against a reference methylation profiles of cell-sorted neutrophils, B cells, monocytes, NK cells, CD4+ T cells and CD8+ T cells, enabling the inference of the cellular composition. Intra-class correlation analyses were performed by conducting a two-way, mixed, consistency analysis comparing samples obtained during response assessment with samples obtained pretreatment using irr (v0.84.1). Statistical significance was defined as a false discovery rate-adjusted p-value < 0.05. Visualizations were generated using ggplot2^111^ (v3.3.5).

### RNA expression and data processing

Transcriptomic analyses was conducted through RNA sequencing, wherein mRNA was extracted utilizing the QIAsymphony system, converted into cDNA and sequenced in a paired-end format on the Illumina NovaSeq6000 at the Amsterdam UMC Core Facility Genomics, generating a dataset comprising 40 million 150 bp-reads. Following that, *in silico* data processing was orchestrated in Snakemake where read quality was assessed using FastQC (v0.11.8) and summarized using MultiQC (v1.0)^92,112^. Raw reads were aligned to the human genome (GRCh38) using the STAR aligner (v2.7.0), with annotations provided by the Ensembl v95 annotation. Post-alignment processing was done in SAMtools (v1.9), followed by read counting using the featureCounts function from the Subread package (v1.6.3)^113–115^. Differential expression analysis (DE) analysis, was carried out within the R statistical environment (v4.3) using the Bioconductor package DESeq2 (v1.38.3). We specifically focused on genes associated with the predictor CpG loci based on the latter’s location in either promoter or enhancer regions. Differentially expressed genes (DEGs) were identified based on their significant differences, defined as those with a Benjamini–Hochberg-adjusted p-value <0.05. Visualization of the results was accomplished using ggplot2 (v3.4.0)^116^.

### Sample size estimation

We based our sample size on an initial pilot experiment and the calculations performed by Tsai and Bell^80^ where we maintained a nominal p-value threshold of 0.05. A pilot experiment using a subset of VDZ treated patients (7 R and 5 NR) indicated on average that the most differentially methylated CpGs presented a mean difference in percentage methylation of 10% when comparing R with NR. We subsequently consulted the power calculations reported by Tsai and Bell^80^, who conducted a case-control epigenome wide simulation study. Assuming an approximately equal number of cases and controls and a mean difference in percentage methylation of at least 10% at a nominal p-value threshold of 0.05, a statistical power of at least 80% would be achieved if we included 40 patients (20 R and 20 NR) per drug. To further eliminate the possibility of being underpowered, we aimed to collect at least 60 patients for the discovery cohort per drug, which would be supplemented by at least 20 patients for the validation cohort.

### Statistical analysis of clinical variables

Baseline characteristics of all included patients were summarized using descriptive statistics. Categorical variables are presented as percentages and continuous variables as median annotated with the interquartile range (IQR). Differences in distribution between responders, non-responders and the different cohorts were assessed using a chi-square test (categorical variables) or Mann-Whitney U (continuous variables). Two-tailed probabilities were used with a p-value ≤0.05 were considered statistically significant. Analyses of clinical data were performed in IBM SPSS statistics (v26). To estimate the probability of a patient responding to either VDZ or USTE after being predicted to be a responder, we calculated the post-test probability using the following formulas:

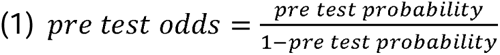

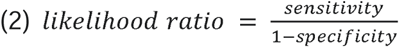

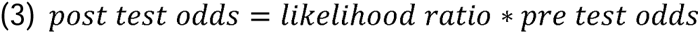

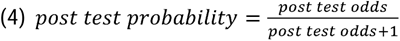

The pre-test probabilities were obtained from the largest VDZ^7^, and USTE^8^ treatment-response studies to date. The sensitivity and specificity were calculated by determining the number of true positives, true negatives, false positives and false negatives when predicting response in the validation cohort.

## Data availability

The raw DNA methylation-(.idat) and gene expression (.fastq.gz) data alongside the de-identified patient metadata as reported on in this study have been published under controlled access for research purposes at the European Genome-phenome Archive (EGA). The DNA methylation data for the VDZ discovery cohort can be found under accession ID: EGAD00010002651. The DNA methylation data for the VDZ validation cohort can be found under accession ID: EGAD00010002652. The DNA methylation data for the USTE discovery cohort can be found under accession ID: EGAD00010002649. The DNA methylation data for the USTE validation cohort can be found under accession ID: EGAD00010002650. The RNA-sequencing data for the VDZ discovery cohort can be found under accession ID: EGAD50000000385. The RNA-sequencing data for the USTE discovery cohort can be found under accession ID: EGAD50000000386.

## Code availability

All Snakemake, bash calls and R scripts have been made available on GitHub and can be found at https://github.com/ND91/HGPRJ0000008_EPICCD_multi_drug.git. The machine learning modeling was performed using proprietary algorithms founded on the same statistical principles as those of gradient boosting, permutation importance, and covered information disentanglement. The code for these techniques is openly available at https://xgboost.ai and https://github.com/JBPereira/CID or https://scikit-learn.org/stable.

## Supplementary Tables

**Supplementary Table 1**: Differential methylation summary statistics of the VDZ-response predictor CpGs. Columns represent the Illumina CpG identifier, the associated HGNC gene symbol, the chromosome, the location on the chromosome (build: hg19), the mean difference in percentage methylation between responders and non-responders, the nominal p-value associated with the mean difference, the intra-class correlation coefficient between pretreatment and during response assessment.

**Supplementary Table 2**: Differential methylation summary statistics of the USTE-response predictor CpGs. Columns represent the Illumina CpG identifier, the mean difference in percentage methylation between responders and non-responders, the nominal p-value associated with the mean difference, the genomic coordinates on the human genome (build: hg19), the annotated gene name.

**Supplementary Table 3**: Clinical characteristics multi-biological failure cohort.

**Supplementary Table 4**: Differential expression summary statistics of the VDZ-response predictor CpGs-associated genes. Columns represent the Ensembl gene ID, the HGNC gene name, the mean difference in log2(fold change) between responders and non-responders and the nominal p-value associated with the mean difference.

**Supplementary Table 5**: Differential expression summary statistics of the USTE-response predictor CpGs-associated genes. Columns represent the Ensembl gene ID, the HGNC gene name, the mean difference in log2(fold change) between responders and non-responders and the nominal p-value associated with the mean difference.

## Acknowledgement

Floris de Voogd for his assistance in evaluation of ultrasound images.

