## Supplementary figures and images for "Peripheral blood DNA methylation signatures predict response to vedolizumab and ustekinumab in adult patients with Crohn’s disease: The EPIC-CD study"

### Supplementary Fig. 1

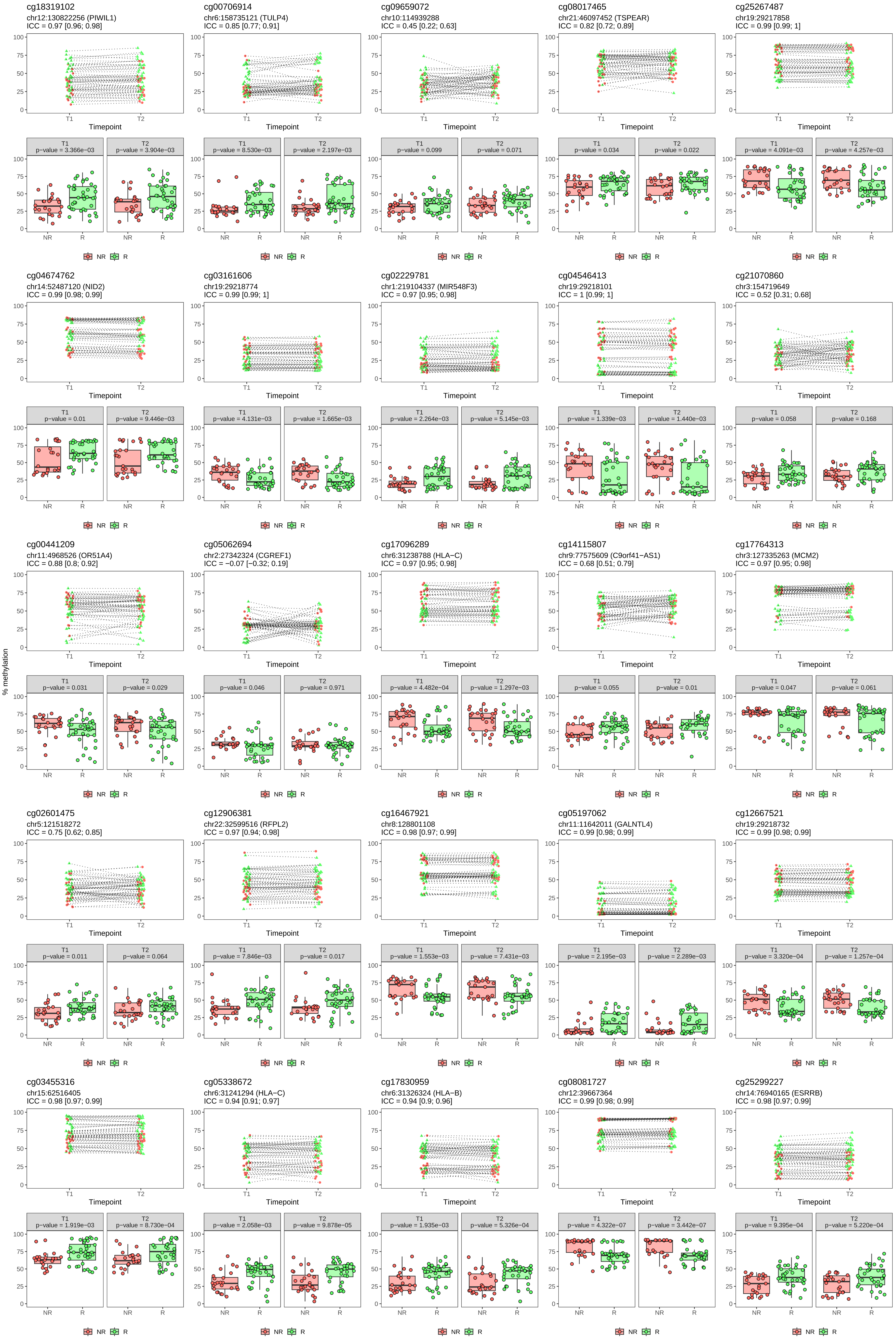

### Supplementary Fig. 2

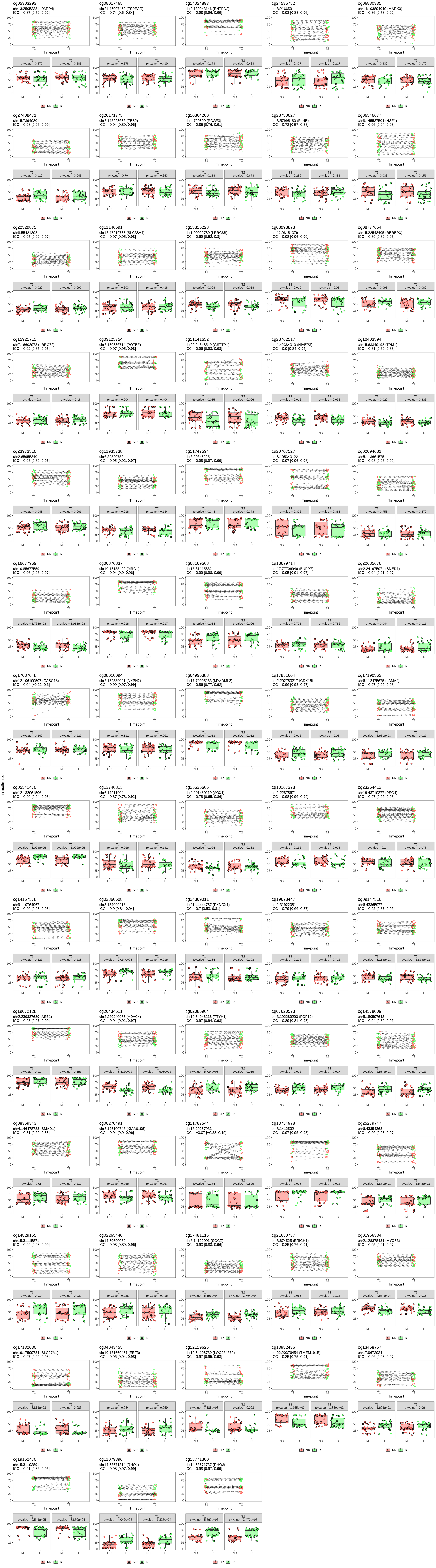

### Supplementary Fig. 3

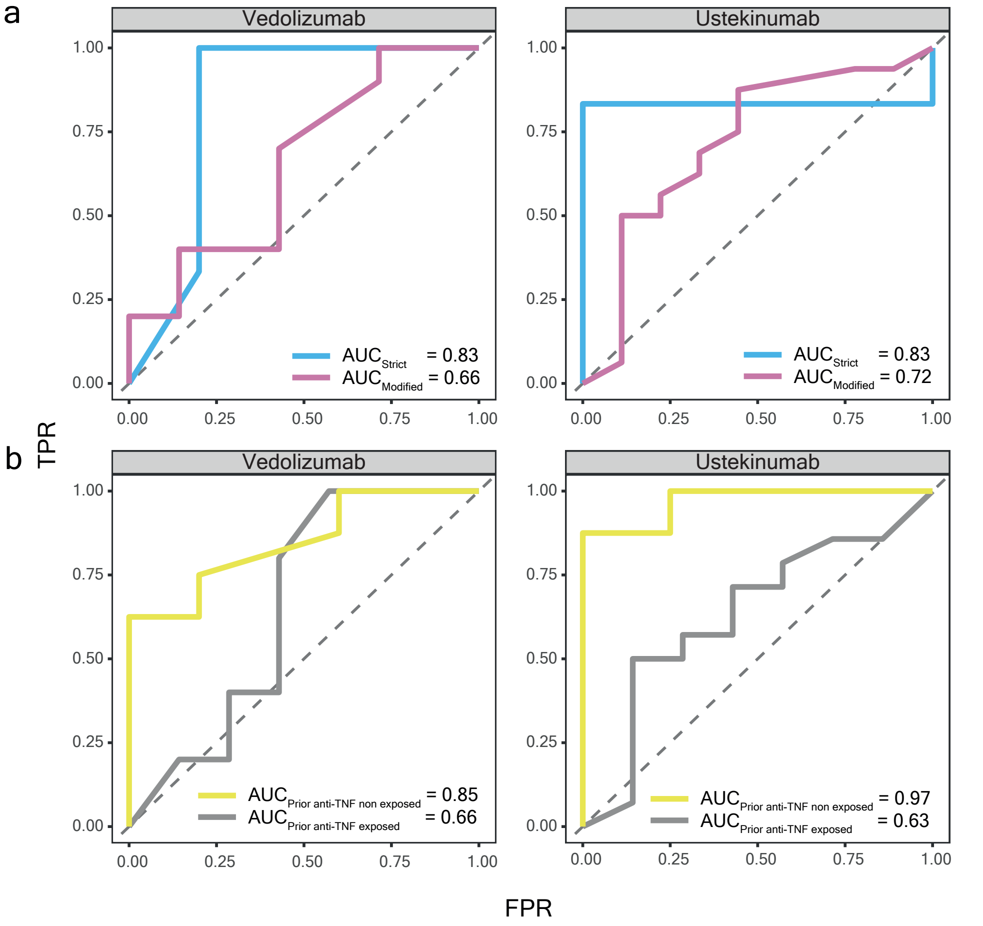

### Supplementary Fig. 4

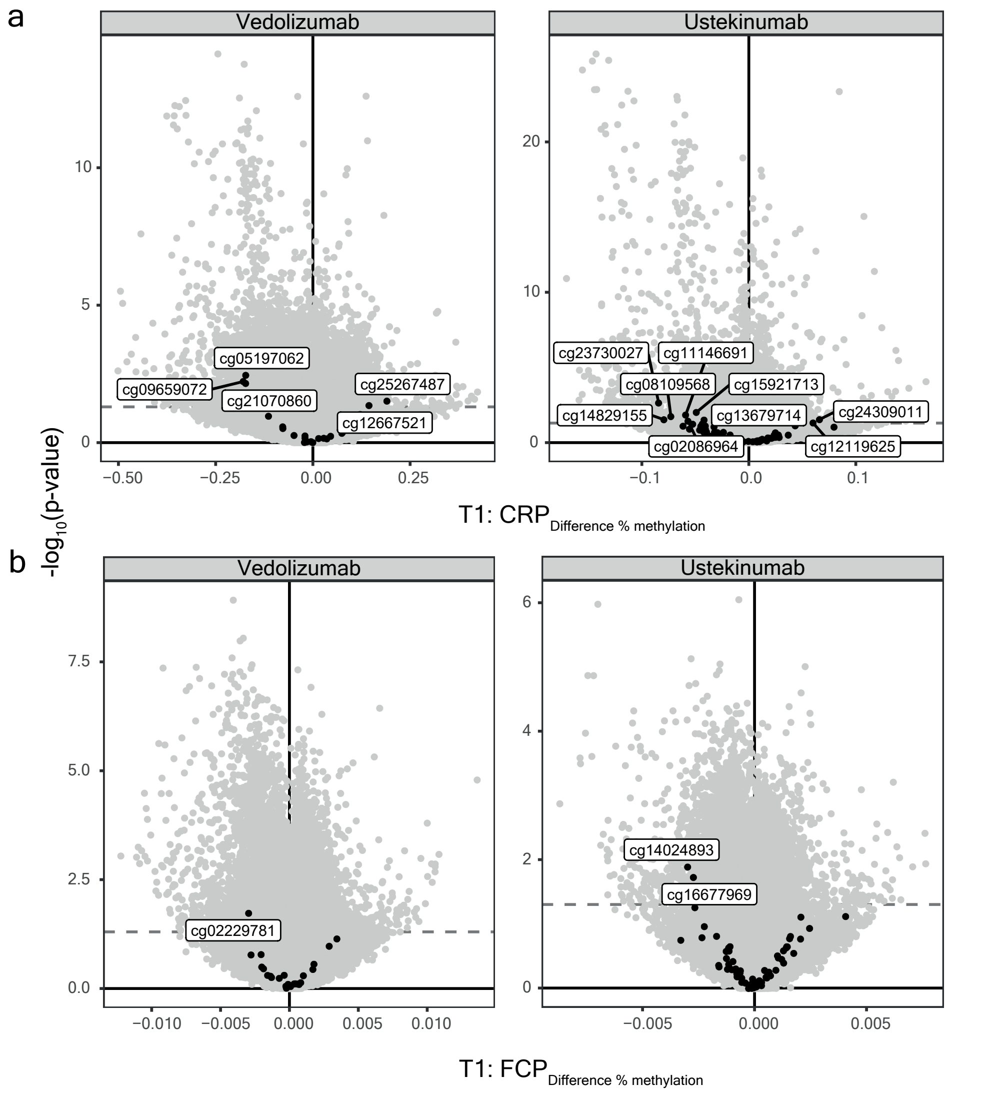
